# COVID-19 Induced Anxiety and Protective Behaviors During COVID-19 Outbreak: Scale Development and Validation

**DOI:** 10.1101/2020.05.05.20050419

**Authors:** Abanoub Riad, Yi Huang, Liping Zheng, Steriani Elavsky

**Affiliations:** Department of Psychology, Faculty of Social Studies, Masaryk University, Joštova 218, 602 00 Brno, Czech Republic; Department of Public Health, Faculty of Medicine, Masaryk University, Kamenice 5, 625 00 Brno, Czech Republic; Department of Psychology, School of Psychology and Cognitive Science, East China Normal University, 3663 N Zhongshan Rd, 200 062 Shanghai, China; Department of Human Movement Studies, Faculty of Education, University of Ostrava, Varenská 40a, 702 00 Ostrava, Czech Republic

**Keywords:** COVID19, Protective Behaviors, Induced Anxiety, Scale Development

## Abstract

**Background:** The outbreak of communicable diseases increases community anxiety levels; however, it demands protective behavioral changes with adjacent awareness of the emerging epidemic. This work aims to develop valid instruments to evaluate COVID-19 induced anxiety, protective behaviors, and knowledge towards COVID-19, and to explore the relationship between the three constructs.

**Methods:** A total sample of 215 university students were recruited to participate in an online self-administered questionnaire. The e-survey consisted of three instruments: COVID-19 Induced Anxiety Scale (CIAS) with 10 items, Protective Behaviors towards COVID-19 Scale (PBCS) with 14 items, and COVID-19 Related Knowledge Scale (CRKS) with 12 items.

**Results:** Item-total analysis and CFA models indicated that CIAS items no. 1, 2, 5, and 8 should be removed to achieve adequate internal consistency (Cronbach’s alpha=0.78) and structural validity. The protective behaviors towards COVID-19 can be estimated from 3 dimensions: Routine Protective Behaviors (RPB), Post-exposure Protective Behaviors (PPB), and Post-exposure Risky Behaviors (PRB). Meanwhile, PBCS showed good internal consistency (Cronbach’s alpha=0.85). Although the sample was unbalanced on gender, gender explained 5% of the variance in protective behaviors with females being more inclined to engage in protective behaviors. Structural Equation Model (SEM) implied that an individual’s COVID-19 related knowledge was associated with the three dimensions of protective behaviors (RPB, PPB and PRB) positively. However, the level of COVID-19 induced anxiety was linked to RPB and PPB positively but negatively to PRB.

**Conclusion:** The 6-item version of CIAS and the 14-item version of PBCS are promising tools for measuring COVID-19 induced anxiety and protective behaviors and can be adopted for future use during early phases of communicable diseases outbreaks. Knowledge is a key indicator for protective behavior; therefore, awareness strategies need to suppress infodemic impact. Severe stress must be monitored during early phases of outbreaks as it significantly increases the probability of risk behavior engagement.

## 1 Introduction

On the last day of 2019, 44 pneumonia cases with unknown etiology were reported to the World Health Organization (WHO) Country Office of China.^1^ This was the first cluster of what would be defined later as coronavirus disease (COVID-19). By January 30th 2020, the WHO had recognized it as a Public Health Emergency of International Concern (PHEIC) due to its rapid spread to 18 countries outside China with recorded cases of human-to-human transmission.^2^ During the following six weeks, more cases were recorded in 114 countries and the death toll reached 4291. Therefore, the WHO declaration of COVID-19 as a pandemic on March 11th 2020, aimed to accelerate the measures undertaken by the member states to avoid an exponential growth of cases which had already occurred in several countries around the world especially in Europe which was recognized as an epicenter of COVID-19 on March 13th 2020.^3, 4^

Non-pharmacologic Interventions (NPIs) are deemed inevitable during the outbreaks of emerging communicable diseases due to a lack of effective drugs and vaccines. The NPIs including public quarantine, social distancing, and case investigation and isolation proved their efficacy in decreasing the production rate in countries that applied aggressive measures e.g. China, Singapore, South Korea, and Taiwan.^5^

The outbreak of communicable diseases is widely perceived as a traumatic event leading to a significant increase in anxiety, depression and fear levels.^6,7^ During the outbreak of the Severe Acute Respiratory Syndrome (SARS)^8^ and the Middle Eastern Respiratory Syndrome (MERS)^9^, the quarantined individuals had more negative emotions like anxiety and anger which are consistent with the findings from isolated mice experiments.^10^ Hu et al. found that the type of quarantine can affect the level of anxiety during the COVID-19 outbreak in China.^11^ Mass quarantine restrictions on non-emergency health services including psychiatric care may adversely affect the access of vulnerable populations to mental health care which cannot be provided by health professionals in isolation units and hospitals due to a lack of specialized training.^7,12^

Posttraumatic stress disorder (PTSD) is a mental health condition characterized by heightened anxiety that can also occur in response to diseases outbreaks. The immediate increase of posttraumatic stress symptoms (PTSS) among patients, healthcare workers, and general population following outbreaks of the SARS, MERS, Ebola, and Zika – had been studied longitudinally in order to evaluate the long-term consequences of these diseases’ outbreaks. SARS survivors with PTSD experienced persistent psychological distress and diminished social functioning in the 4 years after SARS treatment.^13^ Frontline healthcare workers of MERS in South Korea were found experiencing PTSS until 3 years after the outbreak with numbness and sleeping disorders in the high-risk group.^14^ In Italy, 37% of a national survey respondents had PTSS during the 3rd and 4th weeks of lockdown measures – suggesting that monitoring of population’s mental health should be a critical priority during pandemics.^15^

Psychobehavioral surveillance is critical for public health response during communicable disease outbreaks because it informs risk awareness strategies targeting general populations and high-risk groups.^16^ Perceived risk during communicable disease outbreaks motivates people to adopt protective behaviors to reduce any potential hazards of an emerging epidemic.^17^ The relationship between fear and protective behaviors is not linear, but it can be explained by the inverted U-shaped Fear Drive Model of Janis which demonstrates that a moderate level of fear motivates people to adapt protective behavior but when this level is too high or too low, people are more likely to engage in risky behaviors.^18^ Protective behaviors were found to be significantly influenced by the level of knowledge and use of social media during the outbreak of COVID-19 and H1N1 influenza, and to positively impact the epidemic week and viral serial interval.^16,17,19^

Public awareness is a predictor variable which has been associated with both emotions and behaviors during the outbreaks of COVID-19 and H1N1 influenza. Knowledge of the mechanisms of infection transmission and common symptoms are usually found to be sufficient among public, however knowledge of prevention and care-seeking strategies may be distorted by misconceptions and inaccurate information.^20,21^

Infodemic in response to communicable diseases outbreak is an inevitable challenge for public health strategies which occurs as a tsunami of health-related information during early phases of the outbreak – making it challenging for laymen to act properly because of confusing, contradictory, or false information (i.e., fake news), therefore it should be contained in order to escalate behavioral change of public in a predicted manner.^22^

Notably, the definition of “high-risk” groups does not always correlate between epidemiology and psychology. The high-risk groups for infection, disease progression and fatality are predicted to develop more anxious emotions and different patterns of behaviors based on their elevated perceived risk levels. Young adults also form a particular high-risk group during health crises, because they are more susceptible to be influenced by fake news from social media, to have high levels of anxiety and depression, and to engage in risk behaviors.^6,11,23,24^

During communicable disease outbreaks, public health and psychology researchers race against time to assess the early consequences of the emerging phenomena, therefore they typically adopt generic instruments which might not be specific for use in crises settings. To the best of our knowledge, there are no valid instruments to evaluate the induced anxiety, protective behaviors, and public knowledge following communicable disease outbreaks. Therefore, the primary objective of this work was to develop and validate the COVID-19 Induced Anxiety Scale (CIAS), Protective Behaviors towards COVID-19 Scale (PBCS) and COVID-19 Related Knowledge Scale (CRKS). The secondary objective was to evaluate the relationship between communicable disease outbreak induced anxiety, epidemic related knowledge, and protective behavioral tendencies of the adult population.

## 2 Methods

### 2.1 Participants

A self-administered questionnaire with multiple choice items was created in Microsoft Forms (Microsoft Corp. Redmond, WA. 2020). Functionality and user-friendliness of the questionnaire was pre-tested prior to sending it to the participating volunteers by instant messaging applications, WhatsApp (WhatsApp Inc. Menlo Park, CA. 2020) and WeChat (Tencent Holdings Ltd. Shenzhen, China. 2020). University students filled out the questionnaire between March 25th-27th 2020 based on a personal invitation from the study investigators. The URL of the questionnaire was shortened using Bit.ly (Spectrum Equity. Boston, MA. 2020) in order to facilitate its sharing and to enable tracking the visitors. The participants received a gratitude message after completion from the investigators supplied with a factsheet of COVID-19 includes the correct answers of the actual knowledge subscale questions. The study was conducted in accordance with the Declaration of Helsinki^25^ and reported in accordance with the Checklist for Reporting Results of Internet E-Surveys (CHERRIES).^26^ Ethical approval was waived by the Research Ethics Committee (EKV) of Masaryk University because this study did not involve biomedical samples nor did impose greater than minimal risks of information or psychological harms.^27^ An electronic informed consent was obtained from each participant prior to filling the questionnaire. No identifying personal information was collected from the participants upon filling the questionnaire. All the study data were stored in Microsoft Drive in accordance with the General Data Protection Regulation (GDPR).^28^ Participants did not receive any incentives to take part in the study, and they could withdraw at any moment without having to provide justification.

### 2.2 Measures

#### 2.2.1 COVID-19 Induced Anxiety Scale (CIAS)

A 5-point Likert scale with 10 items, where "1" refers to "Totally disagree" and "5" refers to "Totally agree" was developed to evaluate the anxiety induced by the COVID-19 outbreak highlighting the suggested sources of stress and anxious emotions, e.g. “When I or any family member go outside home during this COVID-19 outbreak I feel anxious”. In the theoretical framework, there is only one factor estimated by all items based on the confirmatory factor analysis (CFA) model. The psychometric analysis for the scale was subsequently conducted. (Appendix 1)

#### 2.2.2 Protective Behaviors towards COVID-19 Scale (PBCS)

A 5-point Likert scale with 14 items, where “1” refers to “Not at all like me” and “5” refers to “Just like me” was developed to evaluate people’s protective behaviors against coronavirus infection from 3 dimensions: Routine Protective Behaviors (RPB), Post-exposure Protective Behaviors (PPB), and Post-exposure Risky Behaviors (PRB). Items in RPB are aimed to measure individuals’ protective behaviors in daily life when facing the epidemic, for example, one item in RPB is "I cancel various parties in the event of COVID-19 outbreak immediately". The PPB subscale mainly asks about people’s protective behaviors after the exposure to possible infection. A sample item of PPB is “If I get in contact with someone from COVID-19 outbreak area, I should isolate myself”. Finally, the questions in exam people’s risky behaviors after the possible infective exposure. A sample item of PRB is “If my family member or my friend is in health condition after they come back from outbreak area, there is no need to take protective measures”. According to the previous theory in public survey research, such reversed questions can improve the accuracy of the survey.(55) For the PRB dimension, the item-responses were calculated reversely for the further analysis. In consequence, the higher total scores for each sub-scale and the overall scale refer to the higher quality of protective behaviors. (Appendix 2)

#### 2.2.3 COVID-19 Related Knowledge Scale (CRKS)

A multiple-choice scale of 12 items was developed to assess public awareness of COVID-19 as an emerging communicable disease. Each item has one right option out of four available options. The items were stratified according to 6 major domains: 1) etiology; 2) epidemiological characteristics 3) signs and symptoms; 4) prevention strategies (self-protection); 5) prevention strategies (protection of others); and 6) management measures (while in home quarantine). (Appendix 3)

### 2.3 Statistical Analysis

SPSS 25.0 (SPSS Inc. Chicago, IL. 2020) and the Lavaan package in R were used for statistical analysis.^29^ Three major steps were taken: (1) the item analysis for CIAS and PBCS was conducted based on item-total correlation (54); (2) on the foundation of item analysis, we continued to refine the scales based on the CFA models which aimed to investigate the structural validity of the CIAS and the PBCS; (3) we constructed the SEM model to explore the association of COVID-19 induced anxiety, COVID-19 related knowledge and the protective behaviors in more detail.

## 3 Results

### 3.1 Participants

A total of 215 university students from 17 countries filled out the questionnaire completely. The demographic characteristics of participants are presented in Table 1. The participation rate, defined as the ratio of users who completed the questionnaire / the users who viewed the first page of the survey, was 215/662 (32.5%).

**Table 1.** Demographic Characteristics of Participants

|  | N (Number) | % (Percentage) |
| --- | --- | --- |
| **Gender** |  |  |
| Female | 173 | 80.5 |
| Male | 42 | 19.5 |
| **Age** |  |  |
| 18-20 | 112 | 52.1 |
| 21-25 | 70 | 32.6 |
| 26-30 | 17 | 7.9 |
| 31-35 | 5 | 2.3 |
| 36-40 | 11 | 5.1 |
| **Country** |  |  |
| China | 160 | 74.4 |
| Nigeria | 26 | 21.1 |
| Czechia | 6 | 2.8 |
| Poland | 3 | 1.4 |
| Other | 20 | 9.3 |
| **Academic Level** |  |  |
| Bachelor | 152 | 70.7 |
| Masters | 49 | 22.8 |
| Doctoral | 14 | 6.5 |

### 3.2 Item Analysis

The item analysis was completed using the method of item-total correlation. According to the classical criteria, the item with item-total correlation coefficients below 0.3 should not be accepted.^30^ Thus, for the CIAS, except the first (r=0.17, p=0.01), the second (r=0.10, p=0.13) and the eighth (r=0.19, p<0.01) items, other items were accepted. For the PBCS, the item-total correlations of all questions in scale ranged from 0.34 to 0.73. Therefore, there is no item rejected based on item analysis in the scale.

### 3.3 Confirmatory Factor Analysis (CFA) for COVID-19 Induced Anxiety Scale (CIAS) and Protective Behaviors towards COVID-19 Scale (PBCS)

For the CIAS, we constructed the first CFA model in which the latent level of anxiety was estimated by all remaining 7 items in the original scale after 3 items were rejected by item analysis. However, this model noted the fifth item in CIAS contributed a low factor loading (0.17, less than the recommended 0.4 at the latent construct level).^31^ Thus, following, we continued to delete the item 5 to test the new CFA model, where all factor loadings were above 0.4. Moreover, the model had a good fit (CFI=0.985, RMSEA=0.05, SRMR=0.04, chi-square/df=13.04/9) according to the joint criteria for good model fit (i.e., meeting 3 out of 4 criteria). The 4 specific criteria are (1) CFI>0.9; (2) RMSEA<0.09; (3) SRMR<0.09; (4) chi-square/df <5.^31–33^ The internal consistency was fair (Cronbach’s alpha=0.78).

For the PBCS, we assigned 5 items into the factor of routine protective behaviors (RPB), 6 items into post-exposure protective behaviors (PPB), and 3 items into post-exposure risky behaviors (PRB). All factor loadings were greater than 0.4 and the fit of the CFA model was acceptable (CFI=0.90, RMSEA=0.08, SRMR=0.06, chi-square/df= 179.15/74). The internal consistency of PBCS was good (Cronbach’s alpha=0.85).

### 3.4 Structural Equation Model (SEM) for Induced-Anxiety and Related-Knowledge on Protective Behaviors

Before the SEM, the multiple regression was set up to probe if individual’s demographic information (including gender and academic level) can predict protective behaviors. The results remarked gender accounts for 5% variance of individual’s protective behaviors and that academic level does not explain the protective behaviors. For examining gender’s effect in greater detail, the T-test was done to compare the gender difference in protective behaviors. The outcome inferred females have significantly more protective behaviors than males (t= 3.3, p<0.01). And the COVID-19 induced anxiety and COVID-19 related knowledge account for the variance of protective behaviors by additional 22%.

Theoretically, people’s related knowledge and the anxiety level should influence people’s behaviors towards COVID-19. The Figure 1 depicts the impact of individual’s COVID-19 related knowledge and COVID-19 induced anxiety on the protective behaviors as tested using SEM.

**Figure 1.**
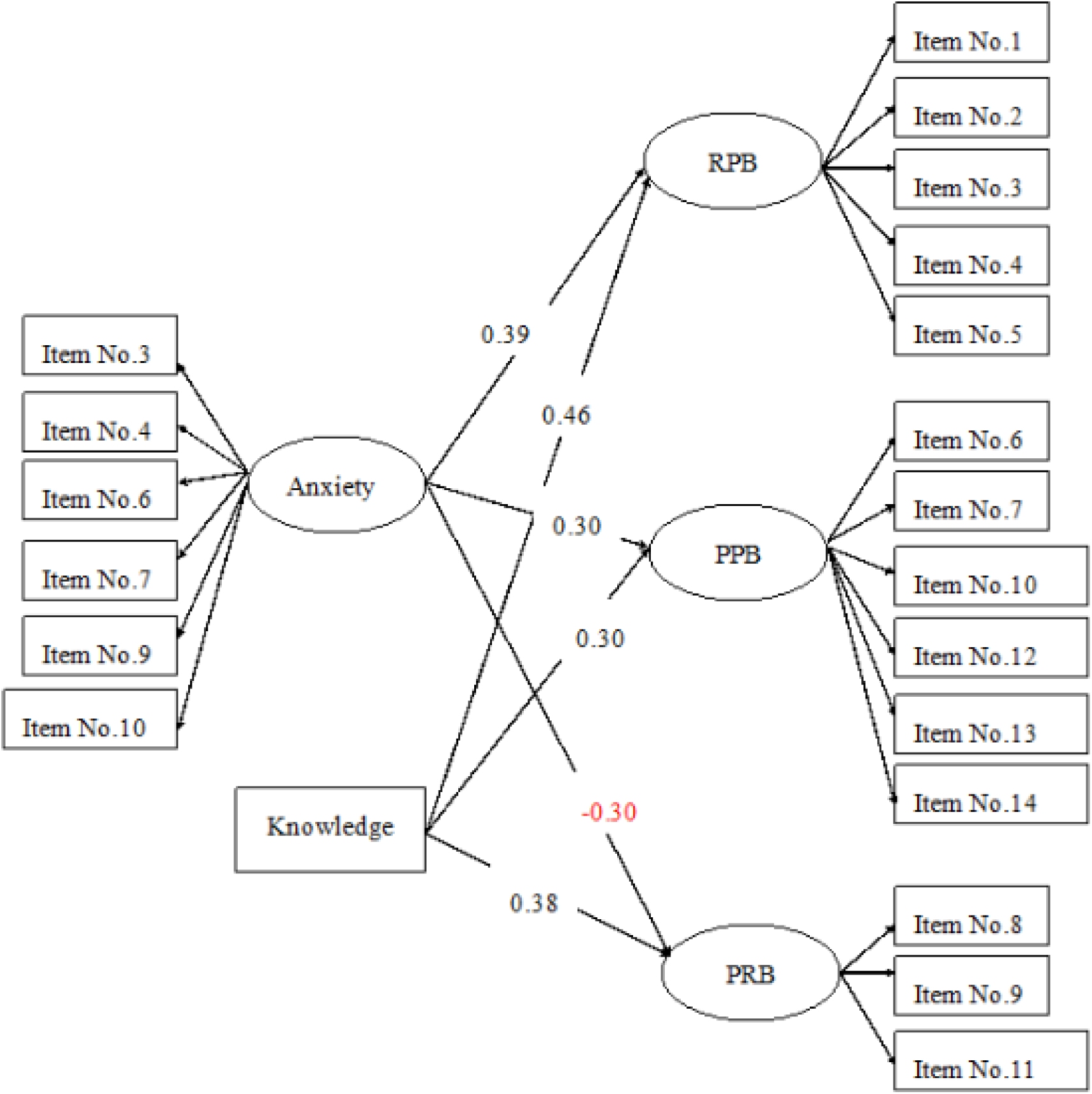
The SEM for describing the impact of COVID-19 related knowledge and COVID-19 induced anxiety on three dimensions of protective behaviors towards COVID-19

The results suggest that COVID-19 related knowledge is positively associated with the three dimensions of protective behaviors towards COVID-19. However, interestingly, the COVID-19 induced anxiety was negatively linked to the dimension of post-exposure risky behaviors (PRB) and positively to routine protective behaviors (RPB) and post-exposure protective behaviors (PPB).

## 4 Discussion

The primary objective of this work was to develop psychometrically sound scales to assess COVID-19 induced anxiety (CIAS), protective behaviors towards COVID-19 (PBCS) and COVID-19 related knowledge. The results indicated that COVID-19 induced anxiety level can be adequately measured by the 6-item version of the CIAS. Communicable diseases outbreaks represent specific health-related crises that may impact people’s emotions in different patterns according to their emerging nature, therefore, the CIAS was designed to cover the potential anxiety sources for general population during COVID-19 outbreak. The CIAS assesses anxiety from specific sources including going outside of the house, disease-related stigma, contracting with people from outbreak areas, getting suspicious clinical symptoms, updates of outbreak data, and resultant death, therefore, it represents a useful tool in measuring level of anxiety specifically related to COVID-19. Interpersonal anxiety-transfer (transmission of anxiety from one person to another) may aggravate during communicable diseases outbreaks through the object-directed social appraisal theory which states that the person becomes significantly influenced by the information picked up from other person’s anxiety expression.^34^

The PBCS can also estimate adequately people’s protective behaviors towards COVID-19 from three aspects: routine protective behaviors (RPB), post-exposure protective behaviors (PPB) and post-exposure risk behaviors (PRB). During the outbreak, any recommended public health measures for individual protection suggested by official health authorities need to be widely perceived and adopted in a timely manner, therefore regular hand-washing, social distancing practice, and face-masks wearing while in public were considered as the RPB of interest in this scale. In contrast to the PPB which include self-isolation and informing local health authorities after coming from abroad, doing home-quarantine and seeking medical advice as soon as suspicious symptoms arise, and reporting neighboring suspicious and confirmed cases, the PRB are about resisting health recommendations by concealing medical history and taking medications without medical advice.

Based on the hierarchical regression model, gender can explain the variation of the protective behaviors while the academic level cannot explain the behaviors. This finding is supported by the t-test which indicated that compared to females, males have lower level of protective behaviors towards COVID-19. Although this finding warrants further corroboration due to the unbalanced and selective nature of this study sample.

Our SEM model implied that an individual’s COVID-19 related knowledge predicts three dimensions of protective behaviors (RPB, PPB and PRB) in a positive linear way. However, for the level of COVID-19 induced anxiety, it only augurs RPB and PPB by positive linear relationship while it is negatively associated with PRB. The gender differences in protective behaviors are consistent with the previous findings which suggested that compared to females, males are more inclined towards risky behaviors for many specific events because females usually perceive more negative possible outcomes than males.^35,36^

The deduction that the level of knowledge towards COVID-19 increases the protective behaviors is consistent with behavioral predictions whereby specific knowledge helps an individual to perform more beneficial behaviors on the knowledge related event. The Health Belief Model argues that individual’s cognitive aspects of health belief, including risk perception and knowledge, can impact the health-related behaviors.^37^ Generally, more comprehensive, and accurate knowledge is linked to more health promoting behaviors. For example, among the elderly population, there is a significant positive correlation between knowledge about Alzheimer’s symptoms and seeking behaviors for professional help.^38^ In college adults, knowledge about AIDS enhances the HIV prevention behaviors.^39^ For schoolchildren, the awareness of the importance of physical exercise promotes their engagement in sport activities.^40^

The interesting phenomenon that the level of COVID-19 induced anxiety not only raises the possibility of routine protective behaviors and post-exposure behaviors, but also enhances the potential post-exposure risky behaviors underscores the complexity with which anxiety impacts behavior. On the one hand, some research indicated that anxiety causes the risk-avoidant decisions and behaviors.^41,42^ The reason behind the decision style can be explained by the theory that anxiety implies the potential threat and so helps people to perceive lower vulnerability to the threat.^43,44^ However, on the other hand, anxiety can have a negative effect, as it could accumulate and make one prone to risk-taking behaviors, especially under circumstances where an individual displays emotion regulation deficits.^45–47^

In conclusion, this study developed and provided initial validation for scales assessing induced anxiety CIAS, protective behavior PBCS, and related knowledge CRKS. These instruments can be rapidly adopted for other communicable diseases during early phase of pandemic outbreaks. Knowledge is a facilitator for protective behaviors, while severe anxiety can be an indicator for risk behaviors during the early phases of outbreak. Therefore, public health strategies need to transmit timely evidence-based health information to the public and to monitor community anxiety and post-traumatic stress symptoms.

## 5 Limitations

First, the participants in the sample were mostly from China and most of them were female. The cultural differences in COVID-19 protective behaviors remain unclear and future studies should evaluate whether the associations among protective behaviors, anxiety and knowledge are stable across gender and cultural contexts.

Second, the knowledge scale was only designed based on professional medical education framework but without explicit psychometric analysis testing if (such as IRT test). Establishing a scales validity is an ongoing process and continues work should ensue to cross-validate the scale in independent samples.

Third, it may be desirable, in addition to assessing actual knowledge, to assess perceived knowledge (e.g., as per Health Belief Model) to evaluate its impact on individual’s protective behaviors towards COVID-19 as well.

Given the limited sample size, future research should continue to evaluate psychometric properties of the scales in more representative samples and further probe the associations among protective behaviors, anxiety and knowledge (actual and perceived).

## 6 Practice Implications

(1) The 6-item version of CIAS and the 14-item version of PBCS are promising tools that can be rapidly adopted to evaluate communicable diseases induced anxiety and protective behaviors during early phase of pandemic outbreaks.

(2) Knowledge is a key indicator for protective behaviors during early phase of the outbreak, therefore public health strategies need to transmit timely evidence-based health information to the public while also highlighting misconceptions circulated by unverified resources.

(3) Severe anxiety in response to communicable diseases outbreak should be monitored as it may lead to risk behaviors which can affect adversely individual’s own health or disease outcomes. Therefore, public health strategies need to monitor community anxiety and post-traumatic stress symptoms.^48^

## Data Availability

The datasets generated and analyzed during the current study are available from the corresponding author on reasonable request.

## 7 Acknowledgements

The authors would like to thank Pascal Bassey MSc. (Masaryk University), Alkauthar Seun Enakele MSc. (Masaryk University), Islam Abdelgawwad BSc. (Damanhour University), Ave Põld DDS. (Charité – Universitätsmedizin Berlin), and each anonymously participating student who contributed to make this work possible.

## 8 Authors Contributions

The first two authors (RIAD A, and HUANG Y) contributed to this work equally. The third author (ZHENG L) contributed to design of the work, while the fourth author (ELAVSKY S) contributed to interpretation of the findings.

## References

1. World Health Organization. Pneumonia of unknown cause – China (2020).

2. World Health Organisation. IHR Emergency Committee on Novel Coronavirus (2019-nCoV) (2020).

3. World Health Organization. WHO Director-General’s opening remarks at the media briefing on COVID-19 - 11 March 2020 (2020).

4. World Health Organization. WHO Director-General’s opening remarks at the media briefing on COVID-19 - 13 March 2020 (2020).

5. Gudi, S. K. & Tiwari, K. K. Preparedness and lessons learned from the novel coronavirus disease. Int. J. Occup. Environ. Medicine 11, 108–112, DOI: 10.34172/ijoem.2020.1977 (2020).

6. Wang, C. et al. Immediate psychological responses and associated factors during the initial stage of the 2019 coronavirus disease (COVID-19) epidemic among the general population in China. Int. J. Environ. Res. Public Heal. 17, 1729, DOI: 10.3390/ijerph17051729 (2020).

7. Li, W. et al. Progression of Mental Health Services during the COVID-19 Outbreak in China, DOI: 10.7150/ijbs.45120 (2020).

8. Hawryluck, L. et al. SARS control and psychological effects of quarantine, Toronto, Canada. Emerg. Infect. Dis. 10, 1206–1212, DOI: 10.3201/eid1007.030703 (2004).

9. Jeong, H. et al. Mental health status of people isolated due to Middle East Respiratory Syndrome. Epidemiol. health 38, e2016048, DOI: 10.4178/epih.e2016048 (2016).

10. V. Heng, M. J. Z. & Smeyne., R. Neurological effects of moving from an enriched environment to social isolation in adult mice (2018).

11. Hu, W., Su, L., Qiao, J., Zhu, J. & Zhou, Y. Countrywide Quarantine Only Mildly Increased Anxiety Level during COVID-19 Outbreak in China. medRxiv; The Prepr. Serv. for Heal. Sci. 2020, 1–20, DOI: 10.1101/2020.04.01.20041186 (2020).

12. Lima, C. K. T. et al. The emotional impact of Coronavirus 2019-nCoV (new Coronavirus disease), DOI: 10.1016/j.psychres.2020.112915 (2020).

13. Hong, X. et al. Posttraumatic stress disorder in convalescent severe acute respiratory syndrome patients: a 4-year follow-up study. Gen. Hosp. Psychiatry 31, 546–554, DOI: 10.1016/j.genhosppsych.2009.06.008 (2009).

14. Lee, S. M., Kang, W. S., Cho, A. R., Kim, T. & Park, J. K. Psychological impact of the 2015 MERS outbreak on hospital workers and quarantined hemodialysis patients. Compr. Psychiatry 87, 123–127, DOI: 10.1016/j.comppsych.2018.10.003 (2018).

15. Rossi, R. et al. COVID-19 pandemic and lockdown measures impact on mental health among the general population in Italy. An N=18147 web-based survey. medRxiv 2020.04.09.20057802, DOI: 10.1101/2020.04.09.20057802 (2020).

16. Wong, L. P. & Sam, I. C. Behavioral responses to the influenza A(H1N1) outbreak in Malaysia. J. Behav. Medicine 34, 23–31, DOI: 10.1007/s10865-010-9283-7 (2011).

17. Oh, S. H., Lee, S. Y. & Han, C. The Effects of Social Media Use on Preventive Behaviors during Infectious Disease Outbreaks: The Mediating Role of Self-relevant Emotions and Public Risk Perception. Heal. Commun. DOI: 10.1080/10410236.2020.1724639 (2020).

18. Janis, I. L. Effects of Fear Arousal on Attitude Change: Recent Developments in Theory and Experimental Research1. Adv. Exp. Soc. Psychol. 3, 166–224, DOI: 10.1016/S0065-2601(08)60344-5 (1967).

19. Lodge, E. K., Schatz, A. M. & Drake, J. M. Protective Population Behavior Change in Outbreaks of Emerging Infectious Disease. bioRxiv 2020.01.27.921536, DOI: 10.1101/2020.01.27.921536 (2020).

20. Moro, M. L. et al. Knowledge, attitudes and practices survey after an outbreak of chikungunya infections. Int. Heal. 2, 223–227, DOI: 10.1016/j.inhe.2010.07.003 (2010).

21. Geldsetzer, P. Use of rapid online surveys to assess people’s perceptions during infectious disease outbreaks: A Cross-sectional Survey on COVID-19, DOI: 10.2196/18790 (2020).

22. Hua, J. & Shaw, R. Corona virus (Covid-19) “infodemic” and emerging issues through a data lens: The case of china. Int. J. Environ. Res. Public Heal. 17, 2309, DOI: 10.3390/IJERPH17072309 (2020).

23. Wang, H.-y. et al. The psychological distress and coping styles in the early stages of the 2019 coronavirus disease (COVID-19) epidemic in the general mainland Chinese population: a web-based survey. medRxiv 2020.03.27.20045807, DOI: 10.1101/2020.03.27.20045807 (2020).

24. Zhong, B. L. et al. Knowledge, attitudes, and practices towards COVID-19 among Chinese residents during the rapid rise period of the COVID-19 outbreak: a quick online cross-sectional survey. Int. journal biological sciences 16, 1745–1752, DOI: 10.7150/ijbs.45221 (2020).

25. (WMA), W. M. A. World Medical Association declaration of Helsinki: Ethical principles for medical research involving human subjects. JAMA – J. Am. Med. Assoc. 310, 2191–2194, DOI: 10.1001/jama.2013.281053 (2013).

26. Eysenbach, G. Improving the quality of web surveys: The Checklist for Reporting Results of Internet E-Surveys (CHERRIES), DOI: 10.2196/jmir.6.3.e34 (2004).

27. Žádost o posouzení – Etická komise pro výzkum | Masarykova univerzita.

28. Proton Technologies AG. General Data Protection Regulation (GDPR) Compliance Guidelines (2020).

29. Rosseel, Y. Lavaan: An R package for structural equation modeling. J. Stat. Softw. 48, 1–36, DOI: 10.18637/jss.v048.i02 (2012).

30. Ferketich, S. Focus on psychometrics. Aspects of item analysis. Res. Nurs. & Heal. 14, 165–168, DOI: 10.1002/nur.4770140211 (1991).

31. Nugent, G. C., Kunz, G. M., Sheridan, S. M., Glover, T. A. & Knoche, L. L. Rural education research in the United States: State of the science and emerging directions (Springer International Publishing, 2016).

32. Gentina, E., Tang, T. L. P. & Gu, Q. Do Parents and Peers Influence Adolescents’ Monetary Intelligence and Consumer Ethics? French and Chinese Adolescents and Behavioral Economics. J. Bus. Ethics 151, 115–140, DOI: 10.1007/s10551-016-3206-7 (2018).

33. Xia, Y. & Yang, Y. RMSEA, CFI, and TLI in structural equation modeling with ordered categorical data: The story they tell depends on the estimation methods. Behav. Res. Methods 51, 409–428, DOI: 10.3758/s13428-018-1055-2 (2019).

34. Parkinson, B. & Simons, G. Worry spreads: Interpersonal transfer of problem-related anxiety, DOI: 10.1080/02699931.2011.651101 (2012).

35. Harris, C. R., Jenkins, M. & Glaser, D. Gender Differences in Risk Assessment: Why do Women Take Fewer Risks than Men? Judgm. Decis. Mak. 1, 48–63 (2006).

36. Lendrem, B. A. D., Lendrem, D. W., Gray, A. & Isaacs, J. D. The Darwin awards: Sex differences in idiotic behaviour. BMJ (Online) 349, DOI: 10.1136/bmj.g7094 (2014).

37. Dobe, M. Health promotion for prevention and control of non-communicable diseases: unfinished agenda. Indian journal public health 56, 180–186, DOI: 10.4103/0019-557X.104199 (2012).

38. Werner, P. Knowledge about symptoms of Alzheimer’s disease: Correlates and relationship to help-seeking behavior. Int. J. Geriatr. Psychiatry 18, 1029–1036, DOI: 10.1002/gps.1011 (2003).

39. DiClemente, R. J., Forrest, K. A. & Mickler, S. College students’ knowledge and attitudes about AIDS and changes in HIV-preventive behaviors. AIDS Educ. Prev. 2, 201–212 (1990).

40. Ferguson, K. J., Yesalis, C. E., Pomrehn, P. R. & Kirkpatrick, M. B. Attitudes, Knowledge, and Beliefs as Predictors of Exercise Intent and Behavior in Schoolchildren. J. Sch. Heal. 59, 112–115, DOI: 10.1111/j.1746-1561.1989.tb04675.x (1989).

41. Maner, J. K. et al. Dispositional anxiety and risk-avoidant decision-making. Pers. Individ. Differ. 42, 665–675, DOI: 10.1016/j.paid.2006.08.016 (2007).

42. Raghunathan, R. & Pham, M. T. All negative moods are not equal: Motivational influences of anxiety and sadnesson decision making. Organ. Behav. Hum. Decis. Process. 79, 56–77, DOI: 10.1006/obhd.1999.2838 (1999).

43. Barlow, D. H. Anxiety and its disorders: the nature and treatment of anxiety and panic (Guilford Press, 2002).

44. Butler, G. & Mathews, A. Anticipatory anxiety and risk perception. Cogn. Ther. Res. 11, 551–565, DOI: 10.1007/BF01183858 (1987).

45. Auerbach, R. P., Abela, J. R. & Ringo Ho, M. H. Responding to symptoms of depression and anxiety: Emotion regulation, neuroticism, and engagement in risky behaviors. Behav. Res. Ther. 45, 2182–2191, DOI: 10.1016/j.brat.2006.11.002 (2007).

46. Cicchetti, D., Ackerman, B. P. & Izard, C. E. Emotions and emotion regulation in developmental psychopathology. Dev. Psychopathol. 7, 1–10, DOI: 10.1017/S0954579400006301 (1995).

47. Cooper, M. L., Agocha, V. B. & Sheldon, M. S. A motivational perspective on risky behaviors: The role of personality and affect regulatory processes. J. Pers. 68, 1059–1088, DOI: 10.1111/1467-6494.00126 (2000).

48. Ray, A., Gulati, K. & Rai, N. Stress, Anxiety, and Immunomodulation: A Pharmacological Analysis. In Vitamins and Hormones, vol. 103, 1–25, DOI: 10.1016/bs.vh.2016.09.007 (Academic Press Inc., 2017).

